# An FDA-approved assay platform can detect biomarkers of neuronal and glial injury in the blood of COVID-19 patients

**DOI:** 10.1101/2024.05.02.24306477

**Authors:** Alexander V. Glushakov, Amy Y. Vittor, Lawrence Lewis, Stacey House, Maggie L. Bartlett, Olena Y. Glushakova, Dan Urbine, Darci R. Smith, Ronald L. Hayes

## Abstract

Employing assays approved by the U.S. Food and Drug Administration (FDA) to assist in detection of brain injury in mild traumatic brain injury (TBI) patients, this study demonstrated that the astroglial protein, glial fibrillary acidic protein (GFAP) and the neuronal protein, ubiquitin C-terminal hydrolase (UCH-L1) were positively associated with age in COVID-19 patients. Controlling for age, UCH-L1 and GFAP were significantly elevated in COVID-19 patients compared to non-COVID-19 controls, and UCH-L1, but not GFAP, was elevated in patients with neurological alterations. Data from this study are also compared to historical data on levels of UCH-L1 and GFAP in brain injured and healthy normal patients. These data support further studies of an FDA approved assay format that could facilitate timely development, validation, and FDA approval of blood tests to detect neuronal and glial cell injuries following infection by SARS-CoV-2. Moreover, appropriately validated blood tests could detect brain injury originating from any systemic pathogen.

**Visual Abstract:** 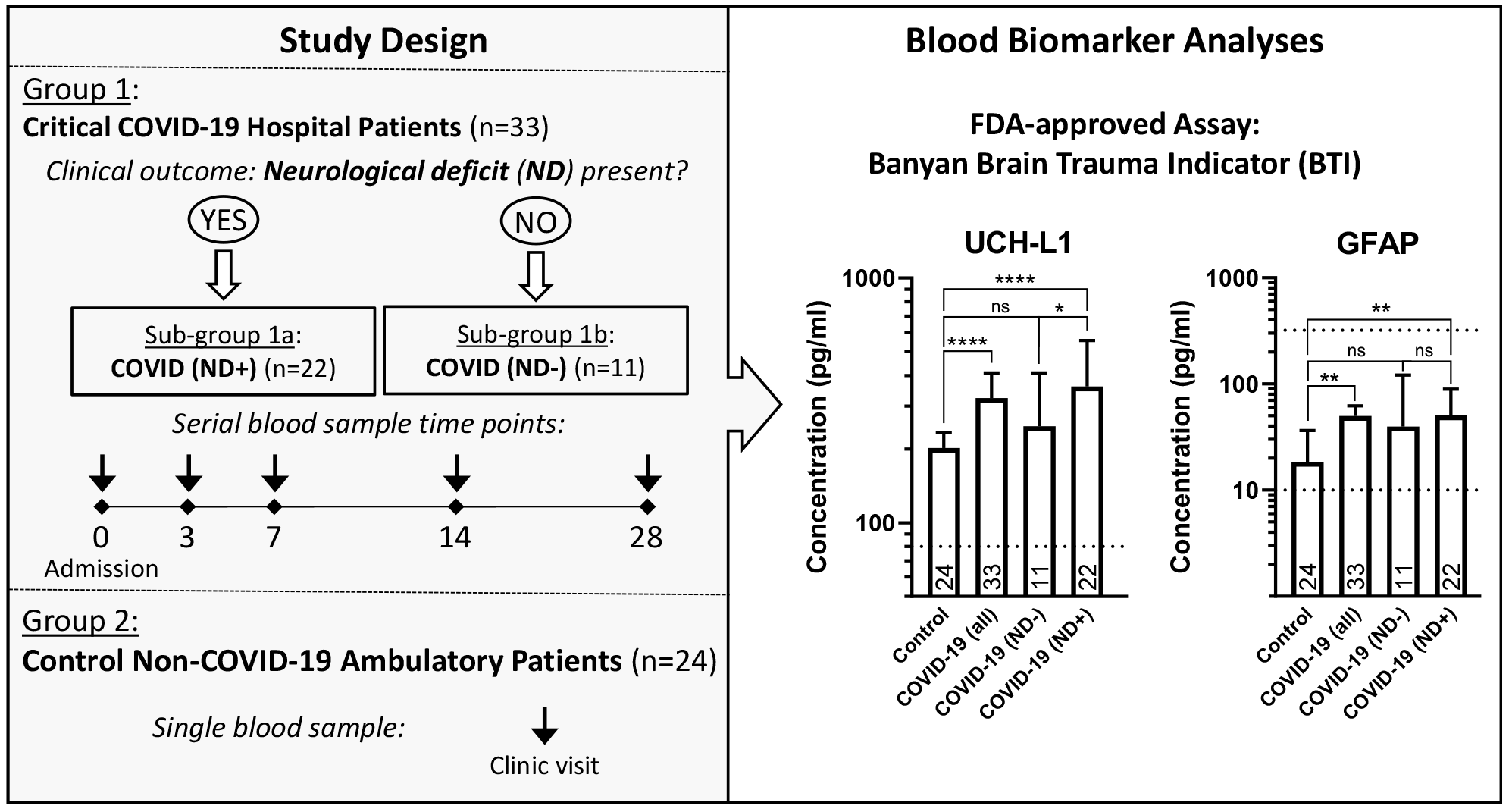

## Introduction

Severe acute respiratory syndrome coronavirus 2 (SARS-CoV-2), the virus that causes coronavirus disease 19 (COVID-19), is now endemic and continues to evolve into potentially more transmissible and pathogenic variants^1, 2^ that challenge vaccine updates.^3^ Future variants could pose greater global health threats due to ongoing antigenic evolution that could lead to new variants that evade immunity.^2^ These mutations have shown increasing escape from neutralizing immunity.^4, 5^ SARS-CoV-2 primarily affects the respiratory system, but it can also have effects on other organs and systems, including the brain. Vaccination before infection confers only partial protection from post-acute sequelae of SARS-CoV-2 (PASC), also known as “long COVID”, a poorly characterized syndrome including a variety of neurological systemic complications.^6, 7^

While COVID-19 is primarily a respiratory illness, patients experience a variety of neurological symptoms resulting from poorly understood mechanisms.^8, 9^ Reported neurologic and psychiatric symptoms of PASC range from headaches, loss of taste and smell, sleep disturbances, cognitive impairment, depression and anxiety. This clinical challenge is complicated by the absence of simple, reliable technologies to diagnose neuropathology in these patients.

COVID-19 patients can experience neuropathology indirectly from systemic complications such as hypoxia or coagulopathy-induced infarcts (for review, see^10^), and autoimmune responses have also been proposed.^11^ The primary proposed mechanisms of neuropathology in acute COVID-19 are systemic inflammation, neuroinflammation, and microvascular injury with leakage of blood products into the parenchyma and microthrombosis.^12, 13^ Some reports maintain that SARS-CoV-2 can directly infect the central nervous system (CNS) but with little evidence of virally mediated injury or inflammation.^14–17^ Acute elevations of fibrinogen and D-dimer relative to C-reactive protein predict cognitive deficits in COVID-19 patients six and 12 months after hospitalization, consistent with a possible contribution of microthrombosis.^18^ Encephalitis is rare, but acute hemorrhagic encephalomyelitis^19^ or an acute disseminated encephalomyelitis^20^ have been reported. Severe COVID-19 is associated with molecular signatures of aging in the human brain,^21^ and cognitive decline can resemble 20 years of aging.^22^ COVID-19 patients also manifest increased incidences of neurological diseases and stroke.^23, 24^

Most COVID-19 transmission occurs asymptomatically.^25^ COVID-19 symptoms persisting for months or longer, and PASC can occur in patients with initially asymptomatic or mild forms of the disease.^26^ Although data vary widely because of varying definitions and poor diagnostic criteria,^27^ PASC, including neurological complications, reportedly affects 6%-69% of patients diagnosed with COVID-19^28–33^ but may vary by SARS-CoV-2 variant.^34, 35^ The potential additional contributions of unexplained post-acute infection syndromes associated with other pathogens^36, 37^ further highlight the need for reliable screening and diagnosis of brain injuries after systemic infections.

Despite appropriate existing technology,^10^ there is still no FDA approved blood test to assist in the diagnosis, prognosis and management of these patients. Increased levels of blood biomarkers of brain injuries have been reported in COVID-19 patients,^38–46^ including glial fibrillary acidic protein (GFAP),^43, 46^ a marker of glial injury, ubiquitin C-terminal hydrolase (UCH- L1),^46^ a marker of neuronal injury and neurofilament light (NfL) and total tau,^42, 43, 46^ markers of axonal damage. GFAP was elevated in a severity dependent manner^43, 46^ and persisted at four months following infection.^43^ UCH-L1 was increased in COVID-19 patients to values greater than seen in non-COVID-19 controls with mild cognitive impairment or Alzheimer’s disease.^46^ NfL was increased in a severity dependent manner,^43, 46^ and improved mortality prediction.^42^ Total tau was increased in a severity dependent manner^46^ and elevations were delayed until four months following infection.^43^ Elevations of GFAP, but not NfL, have also been reported in PASC patients, although GFAP elevations were not associated with subjective symptom reports.^47^

In this study we employed an assay measuring levels of GFAP and the neuronal protein ubiquitin C-terminal hydrolase (UCH-L1) developed by Banyan Biomarkers, Inc. and approved by the FDA to assist in detection of brain injury in mild traumatic brain injury (TBI) patients.^48^ The assay format can also be used to predict functional recovery after TBI.^49, 50^ As we have pointed out elsewhere,^10^ the use of an FDA approved assay platform significantly facilitates the development, validation and ultimate FDA approval of blood tests to detect brain pathology following infection by SARS-CoV-2. Moreover, appropriately validated blood tests could detect brain injury originating from any systemic pathogen.^10^

## Methods

### Study populations and sample collection procedures

The primary study was performed on a subgroup of subjects who presented to the Barnes-Jewish Hospital Emergency Department (ED) in St. Louis during a one-month period from March 27, 2020 through April 27, 2020 with signs or symptoms of COVID-19 who were prospectively enrolled in the study and consented to having serial blood samples obtained during their hospitalization and stored for future analysis. We analyzed 71 serum samples from 33 unique patients with signs or symptoms of COVID-19 and a positive RT-PCR test for SARS-CoV-2 at some point during their hospitalization. Serum samples were obtained, when feasible, at the time of presentation and day 3, day 7, day 14, and day 28 for patients still in the hospital. The blood sample was spun down to serum and stored at -80°C. The serum samples were sent to the Naval Medical Research Command, Biological Defense Research Directorate, Ft. Detrick, MD for analyses. We compared the serum concentration of UCH-L1 and GFAP in patients with neurologic signs or symptoms to those without. We further stratified results by the day on which the serum sample was obtained.

Our study population consisted of 24 men and 9 women, ranging in age from 27 years to 88 years, with a mean age of 64 ± 14 years. Eleven patients had no neurologic signs or symptoms, while the remainder, 22, had signs/symptoms ranging from anosmia to altered mental status and unresponsiveness (see **Supplemental Table 1**). Overall, the prevalence of neurological deficits in COVID-19 positive subjects in males and females were 71% and 56%, respectively, but the difference was not statistically significant (P = 0.4376, Fisher’s exact test). Twenty-five patients (including all 11 without neurologic signs or symptoms) did not have neuro-imaging.

The findings of the imaging on the 15 patients who underwent it, the time of sampling, and the serum concentration of UCH-L1 and GFAP stratified by neurologic and imaging findings is available in the **Supplemental Table 1**.

Twenty-four non-COVID-19 controls with equal gender proportion (not significantly different from the COVID-19 group, P=0.1, Fisher’s exact test) and ranging in age from 23 years to 84 years, with an average age of 50 ± 19 years were obtained from the University of Florida (UF) Clinical and Translational Science Institute Biorepository. These specimens were originally obtained from the UF Health System’s outpatient population prior to 2019, and were therefore not exposed to SARS-CoV-2 and did not have COVID-19 symptoms. Some of the non-COVID-19 controls carried cancer or benign tumor diagnosis, but specimens from individuals with conditions including brain trauma, cerebral vascular events, or dementia were excluded. Samples were stored and shipped for analyses as with Barnes-Jewish samples.

### Brain biomarker measurements

All de-identified sera samples were evaluated with the Banyan Brain Trauma Indicator (BTI) assay according to manufacturer’s guidelines. For primary data analyses, the biomarker concentrations were adjusted to reportable ranges of the FDA approved BTI assay as described elsewhere,^48^ whereas for additional analyses to further explore associations of biomarker levels with COVID-19 diagnosis, neurological outcomes and demographic variables using quantile regression analysis, actual device readings were used in samples with UCH-L1 or GFAP concentration below the reportable BTI assay ranges, if available, or analytically determined actual concentration in the samples with biomarker concentration above the reportable ranges (see **Supplemental Table 1**). If the concentration was above the upper limit of quantitation for the assay in selected samples (2 UCH-L1 samples and 2 GFAP samples of a total of 95 samples tested in each assay), then a dilution of the serum was made, and the sample was re-run and the concentration was reported incorporating the dilution factor. Nine of 65 (13%) and 6 of 24 (25%) of GFAP samples assessed in COVID-19 group and non-COVID-19 control group, respectively, were below the lower limit of quantitation (LLOQ), whereas no UCH-L1 samples felt below the LLOQ. The samples that were below the LLOQ for the GFAP assay are indicated as <10.

### Statistical analyses

Demographic data were tested for normality, if appropriate (e.g., age), using the Shapiro–Wilk test and proportions of males and females between COVID-19 and non-COVID-19 control groups as well as proportions of neurological deficit prevalence between genders were assessed using Fisher’s exact test (GraphPad Prism 8, GraphPad Software, Boston, MA).

Since the main focus of this study was to explore the ability of the FDA-approved Banyan BTI assay to detect biomarkers of CNS injury in critical COVID-19 patients, primary data analyses (the data described in **Figures 1** and **2**) reflected reportable ranges for its intended clinical application (see also^48^). The data were analyzed using GraphPad Prism 8 software. Details of statistical analyses are included in the figure legends or in the text of the article. In general, to handle the analyses of the data of the serial sample collection with missing time points, we applied a mixed-model statistical approach with one-way analysis and multiple comparison between groups, which assumes fixed main factors in random study subjects (see also^51, 52^).

**Figure 1:**
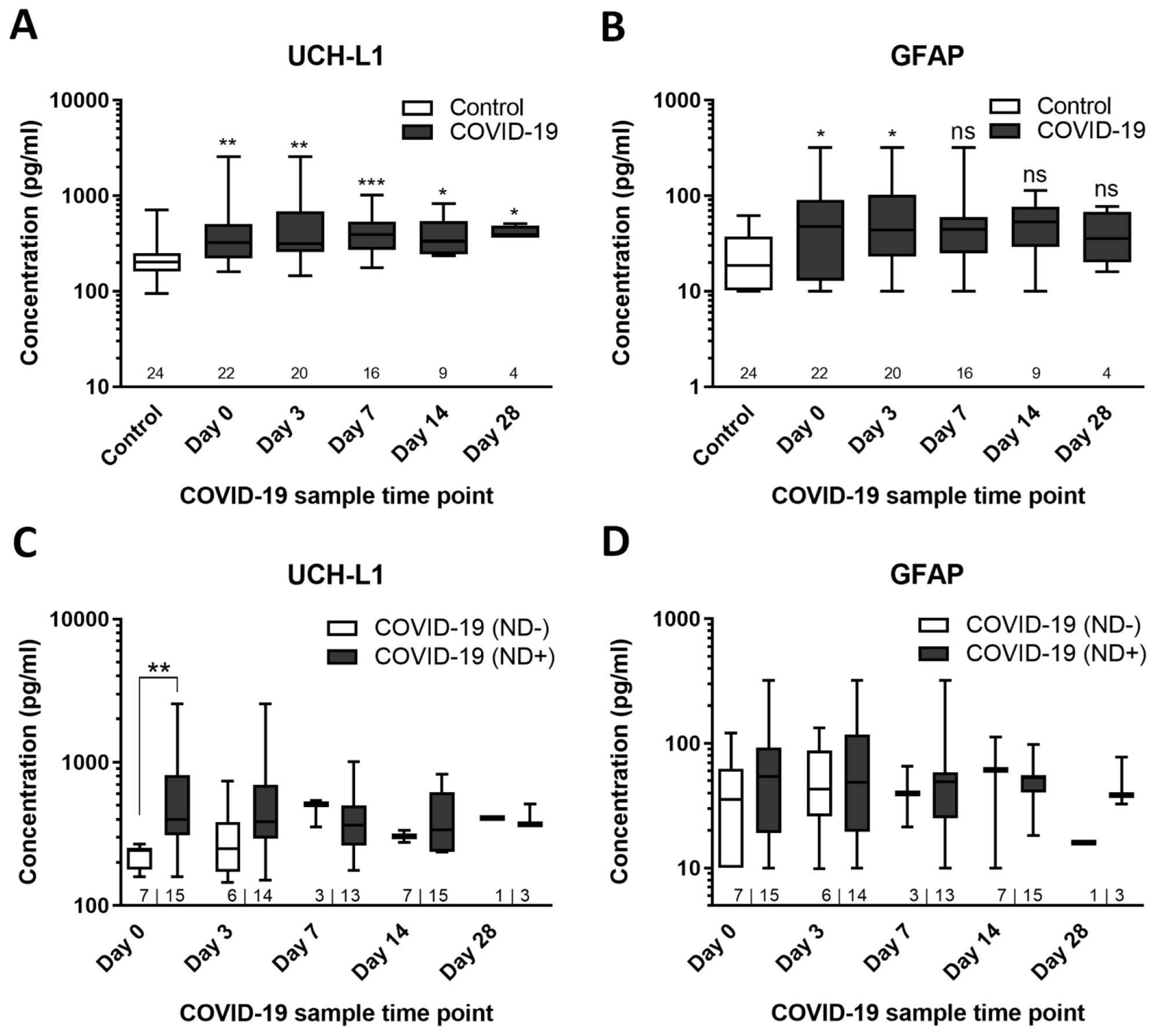
UCH-L1 and GFAP levels in COVID-19 positive patients, with and without neurological changes, vs. non-COVID-19 control subjects. **A** and **B**, The UCH-L1 and GFAP levels in COVID-positive patients at different time points compared to non-COVID-19 controls. Box and whiskers graphs represent the median, min-to-max and interquartile ranges. The numbers above the X axes represent sample sizes per group per time point. Asterisks indicate statistical significance of comparison biomarker values at corresponding time points using Kruskal-Wallis test with Dunn’s post-hoc test vs. non-COVID-19 control group (*P &lt; 0.05, **P &lt; 0.01 and ***P &lt; 0.001). “ns” indicates not statistically different (P > 0.05). There were no statistically significant differences between biomarker values between time points in the COVID-19 positive subjects. **C** and **D**, Time courses of UCH-L1 and GFAP levels in COVID-19 positive subjects grouped based on the presence or absence neurological changes. Box and whiskers graphs represent the median, and min-to-max and interquartile ranges. The numbers above the X axes represent sample sizes per group per time point. Asterisks indicate statistical significance of comparison biomarker values of samples from subjects with and without neurological changes at corresponding time points using Kruskal-Wallis test with Dunn’s post-hoc test (**P&lt;0.01). Other biomarker values were not statistically significantly different.

**Figure 2:**
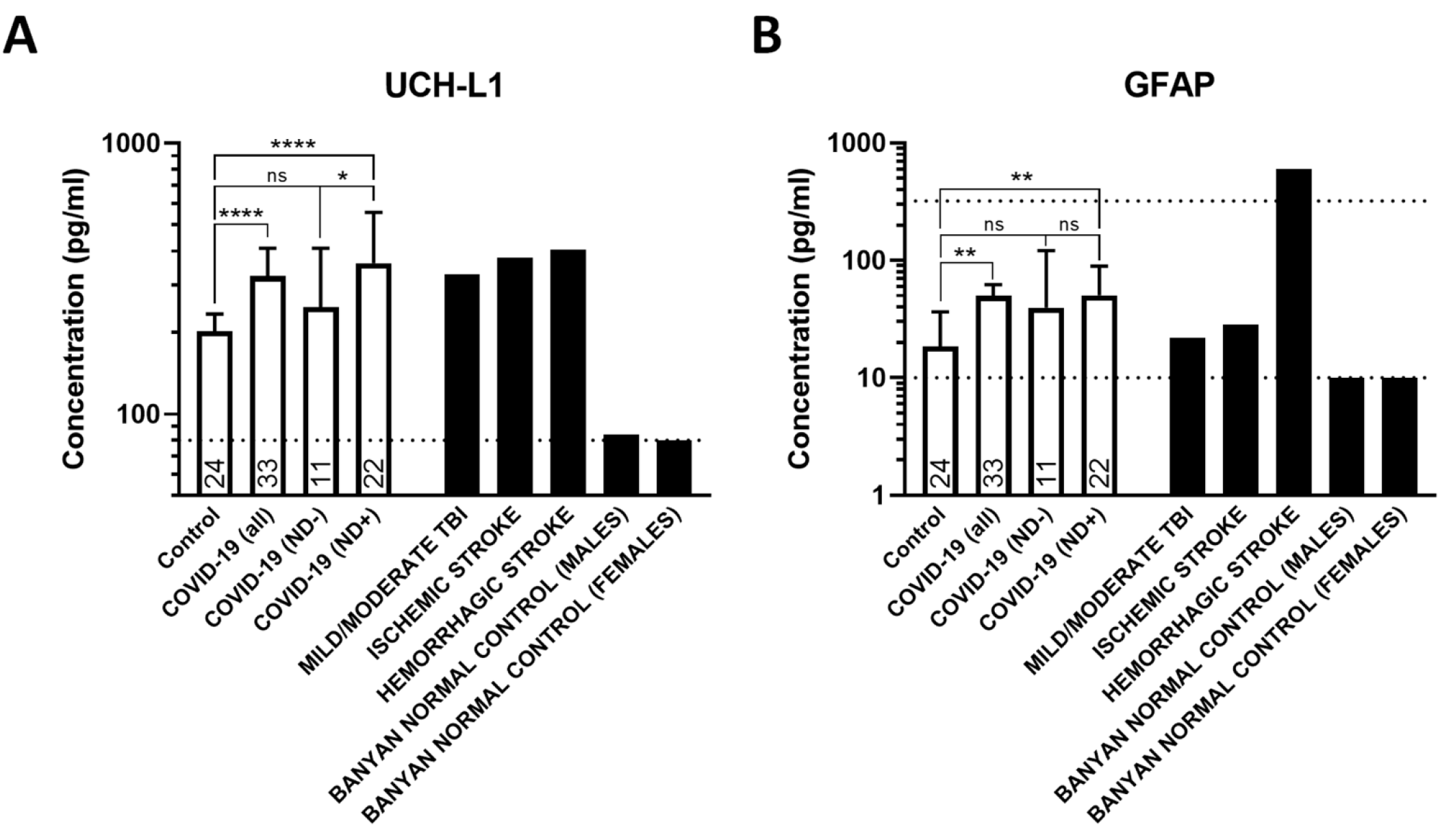
UCH-L1 and GFAP levels in COVID-19 positive patients vs. non-COVID-19 controls and historical data on TBI and stroke patients and healthy populations. **A** and **B**, UCH-L1 and GFAP levels in COVID-19 positive patients compared to non-COVID-19 controls from this study (white filled bars) and historical cutoff values in neurological injuries^48, 53^ and healthy control populations shown for comparison (black filled bars), respectively. COVID-19 (all), COVID-19 (ND+) and COVID-19 (ND+) denote group of all COVID-19 positive subjects and subgroups of COVID-19 positive subjects with and without neurological changes, respectively. Dotted lines indicate lower and upper reportable range values of the BTI assay (i.e., 80–2560 pg/mL for the UCH-L1 assay and 10–320 pg/mL for the GFAP), if applicable. For groups of non-COVID-19 control and COVID-19 positive subjects, bars represent median and “error” bars represent 95% CI. Values for each bar in each COVID-19 positive subjects were calculated as median biomarker levels of the serial samples, if applicable. The numbers above the X axes represent sample sizes per group. Asterisks indicate statistical significance of comparison biomarker values between corresponding groups (*P &lt; 0.05, **P &lt; 0.01 and ****P &lt; 0.0001) and “ns” indicates not statistically different (P > 0.05). For comparison two groups [i.e., non-COVID-19 control vs. COVID-19 (all)], two-tailed Mann Whitney test was used, whereas for comparison multiple groups [i.e., non-COVID-19 control vs. COVID-19 (ND+) vs. COVID-19 (ND+)], Kruskal-Wallis test with Dunn’s multiple comparison post-hoc test was used.

In additional analyses, to evaluate the association between UCH-L1 or GFAP and COVID-19 positivity, neurological symptoms, age and sex, quantile regression estimating the conditional median was performed since not all conditions of linear regression were met. To perform the aforementioned quantile regression analyses, average UCH-L1 and GFAP values for days 0, 3, 7, 14 and 28 were determined for the COVID-19 positive individuals. UCH-L1 and GFAP values were assessed at a single time point for the non-COVID-19 control individuals. Data were analyzed in Stata version 14.2 (StataCorp LLC, College Station, TX).

P values less than 0.05 were considered significant. Non-continuous and mixed data were presented as median and min-to-max and interquartile ranges or 95% confidence interval (CI). Numerical descriptive statistic values of normally distributed data (e.g., age) were reported as mean ± SD.

## Results

Figures 1A and **B** present the time courses of UCH-L1 and GFAP levels in COVID-19 positive patients vs. non-COVID-19 controls. Levels of UCH-L1 were elevated throughout the 28-day sample collection period. Levels of GFAP were more variable and elevated only on days 0, 3 and 14. The UCH-L1 and GFAP concentrations were not significantly different between time points in the COVID-19 positive subjects. Figures 1C and **D** present the time courses of UCH- L1 and GFAP in COVID-19 positive patients with vs. without neurological changes. Only UCH- L1 showed significant elevations in neurologically altered patients on the day of study enrollment (day 0).

In order to add context to the observed magnitude of changes in biomarkers, Figure 2 compares biomarker levels in samples collected in the current study to levels observed in patients experiencing a TBI who are at risk for intracranial bleeding,^48^ in patients who have had hemorrhagic or ischemic stroke^53^ and in a healthy population. Necessary for valid comparisons, all assays were conducted using the same FDA approved assay format.^54^ Median levels of UCH-L1 in the COVID-19 group calculated for each subject from the available time points from day 0 through day 28 were similar to levels in stroke and TBI patients. Levels of GFAP in COVID-19 positive patients were only exceeded by levels in hemorrhagic stroke patients. The levels of UCH-L1 and GFAP in non-COVID-19 controls were markedly elevated compared to community healthy controls, both males and females. Statistical analyses presented in Figure 2 further demonstrate significant differences between both UCH-L1 and GFAP biomarker values in non-COVID-19 controls and median biomarker values (P < 0.001 and P < 0.01 for UCH-L1 and GFAP, respectively). Comparison between non-COVID-19 control group with a sub-group of COVID-19 positive subjects with neurological deficits demonstrate significant differences, as well (P < 0.001 and P < 0.05 for UCH-L1 and GFAP, respectively), whereas there were no significant differences between non-COVID-19 control group and a sub-group of COVID-19 positive subjects without neurological deficits. Interestingly, there was statistically significant differences between median values of UCH-L1 concentrations in sub-groups of COVID-19 positive subjects with and without neurological deficits (P < 0.05), but not for GFAP concentrations.

Importantly, additional statistical analyses comparing nadir and peak UCH-L1 concentrations calculated for individual subjects in COVID-19 group with the biomarker values in non-COVID- 19 controls yielded similar results to those comparing of median values shown in Figure 2.

When compared to non-COVID-19 controls, both nadir and peak UCH-L1 values were statistically different in COVID-19 positive subjects (P < 0.01 and P < 0.001 for nadir and peak values, respectively) and in the subgroup of COVID-19 positive subjects with neurological deficits (P < 0.01 and P < 0.001 for nadir and peak values, respectively), whereas only peak GFAP values were statistically different (P < 0.01 for both comparisons). When controlling for gender using the same statistical analyses, median and peak UCH-L1 values were significantly different in COVID-19 positive subjects regardless of neurological status (P < 0.05 and P < 0.01 for men, and P < 0.05 for women, respectively) as well as in sub-groups of COVID-19 positive subjects with neurological deficits (P < 0.01 for men and P < 0.05 for women, respectively) as compared to the biomarker concentrations in non-COVID control subjects. The median and peak GFAP values in women were also significantly different in COVID-19 positive subjects regardless of neurological status (P < 0.05) as well as the nadir, median and peak GFAP values in sub-groups of COVID-19 positive subjects with neurological deficits (P < 0.05) as comparing to the biomarker concentrations in non-COVID control subjects, whereas only peak GFAP values in male COVID-19 positive subjects were significantly different compared to non-COVID controls (P < 0.05).

Quantile regression analysis demonstrated that UCH-L1 was associated with age (in years) in men (β=5.67, 95% CI 0.94, 10.4, p<0.05), but not in women. Controlling for age and sex, UCH- L1 concentrations were significantly higher (95% CI 68.81, 277.68, p<0.01) in COVID-19 patients presenting with neurological complications compared to non-COVID-19 controls.

However, no statistically significant association was observed for UCH-L1 in COVID-19 positive patients without neurological complications compared to non-COVID-19 controls.

Similar to UCH-L1, the median GFAP value was associated with age (in years) in men (β=0.96, 95% CI 0.30, 1.63, p<0.01) but not in women. Controlling for age and sex, no statistically significant association was observed for GFAP in COVID-19 positive patients with or without neurological complications compared to non-COVID-19 controls.

## Discussion

Although this retrospective study employs opportunistic samples and modest sample sizes, the research employs an FDA cleared assay format and documents the feasibility of future research examining an accelerated pathway to an FDA approved diagnostic assay for rapid diagnosis of neuropathology in COVID-19 patients. In addition to UCH-L1 and GFAP, future studies should consider additional biomarkers including NfL. Even mild respiratory COVID-19 can result in loss of myelin, oligodendrocytes and myelinated axons,^39^ and NfL is elevated in COVID-19 patients.^42, 43^ Previous studies have demonstrated that SARS-CoV-2 directly infects brain astrocytes.^15, 16, 55^ Our study did not detect differences in GFAP between patients with and without neurological alterations, although GFAP was elevated in COVID-19 patients vs. non-COVID-19 controls. Future research should more thoroughly examine astroglial involvement in COVID-19 patients.

As reviewed above, a predominance of studies employing different COVID-19 patient cohorts, times of biomarker assessments and assay formats have reported increased levels of brain injury biomarkers including GFAP and UCH-L1. Our limited data precludes examinations of relationships of biomarker changes to the extent of brain injuries or magnitudes/durations of neurological deficits, important goals of future studies. Moreover, comparisons to historical data in Figure 2 using the same FDA approved brain injury format suggest the possibility of clinically important brain injury in some COVID-19 patients. The statistical differences in the UCH-L1 levels at early time points (e.g., day 0 in our study) in COVID-19 positive subjects even with and without neurological alterations suggests the utility of this biomarker for prediction of neurological complication in COVID-19.^18^ Further, the statistical differences in nadir, median and peak UCH-L1 and GFAP concentrations from serial sampling also suggest a possible extended application of this FDA approved assay to provide evidence-based clinical decisions to improve COVID-19 patient management. Elevations of UCH-L1 and GFAP in non-COVID-19 controls used in this study, which might be potentially associated with a concomitant medical condition (e.g., cancer), over levels seen in healthy population also merits further study.

Since the same assays used in this study could potentially detect neuronal and glial cell injury from infections by any systemic pathogens, this research can provide urgently needed technology to diagnose currently unrecognized increased risks for neurodegenerative diseases from numerous other infections.^36, 37, 56^ SARS-CoV-2 can also potentially result in opportunistic infections by other neurotropic viruses such as the human herpes virus which has been reported in 79% of severe COVID-19 patients.^57^ Thus, it could be important to screen for other neurotropic pathogens in COVID-19 patients with neurological deficits.

## Transparency, Rigor, and Reproducibility Summary

Samples were collected under institutional review board (IRB) approval from their respective institutions: Barnes-Jewish Hospital ED-in St. Louis, IRB#202007018 utilizing samples collected in a COVID-19 repository approved under IRB#202003085; University of Florida, utilizing samples collected by Clinical and Translational Science Institute Biorepository under IRB#202001993. All samples were de-identified and sent to the Naval Medical Research Command (NMRC), Biological Defense Research Directorate, Ft. Detrick, MD. The protocol for this study was approved by the NMRC Institutional Review Board in compliance with all applicable federal regulations governing the protection of human subjects. This study is not a subject of clinical trial registration requirements. Biomarker analyses in de-identified samples were performed using an FDA-approved BTI assay directly supplied from the manufacturer. The assay was performed by an investigator blinded to patients’ clinical characteristics including the presence or absence neurological manifestations. The reportable ranges for this assay are 80– 2560 pg/mL for the UCH-L1 assay and 10–320 pg/mL for the GFAP assay. The assay results, patient clinical characteristics and selected unidentified demographic data are provided as supplemental materials. The further details on human sample collection, technical assay procedures, statistical analyses are described in the corresponding method sections.

## Authors’ Contributions

The authors confirm their contribution to the paper as follows: Conceptualization: A.V. Glushakov, D.R. Smith, R.L. Hayes; Formal analysis: A.V. Glushakov, A.Y. Vittor; O.Y. Glushakova; Funding acquisition: D.R. Smith; Investigation: L. Lewis, S. House, M.L. Bartlett, D.R. Smith; Resources: A.Y. Vittor; L. Lewis, S. House, D.R. Smith; Visualization: A.V. Glushakov; Writing - Original Draft: A.V. Glushakov, R.L. Hayes; Writing - Review & Editing: A.V. Glushakov, A.Y. Vittor; L. Lewis, S. House, M.L. Bartlett, O.Y. Glushakova, D. Urbine, D.R. Smith, R.L. Hayes. All authors reviewed the results and approved the final version of the manuscript.

## Data Availability

The assay results, patient clinical characteristics and selected unidentified demographic data are provided as supplemental materials.

## References

1. Kimura, I., Yamasoba, D., Tamura, T., Nao, N., Suzuki, T., Oda, Y., Mitoma, S., Ito, J., Nasser, H., Zahradnik, J., Uriu, K., Fujita, S., Kosugi, Y., Wang, L., Tsuda, M., Kishimoto, M., Ito, H., Suzuki, R., Shimizu, R., Begum, M.M., Yoshimatsu, K., Kimura, K.T., Sasaki, J., Sasaki-Tabata, K., Yamamoto, Y., Nagamoto, T., Kanamune, J., Kobiyama, K., Asakura, H., Nagashima, M., Sadamasu, K., Yoshimura, K., Shirakawa, K., Takaori-Kondo, A., Kuramochi, J., Schreiber, G., Ishii, K.J., Genotype to Phenotype Japan, C., Hashiguchi, T., Ikeda, T., Saito, A., Fukuhara, T., Tanaka, S., Matsuno, K. and Sato, K. (2022). Virological characteristics of the SARS-CoV-2 Omicron BA.2 subvariants, including BA.4 and BA.5. Cell 185, 3992–4007 e3916.

2. Markov, P.V., Katzourakis, A. and Stilianakis, N.I. (2022). Antigenic evolution will lead to new SARS-CoV-2 variants with unpredictable severity. Nature reviews. Microbiology 20, 251–252.

3. Callaway, E. (2022). Fast-evolving COVID variants complicate vaccine updates. Nature 607, 18–19.

4. Khan, K., Karim, F., Ganga, Y., Bernstein, M., Jule, Z., Reedoy, K., Cele, S., Lustig, G., Amoako, D., Wolter, N., Samsunder, N., Sivro, A., San, J.E., Giandhari, J., Tegally, H., Pillay, S., Naidoo, Y., Mazibuko, M., Miya, Y., Ngcobo, N., Manickchund, N., Magula, N., Karim, Q.A., von Gottberg, A., Abdool Karim, S.S., Hanekom, W., Gosnell, B.I., Team, C.-K., Lessells, R.J., de Oliveira, T., Moosa, M.S. and Sigal, A. (2022). Omicron BA.4/BA.5 escape neutralizing immunity elicited by BA.1 infection. Nat Commun 13, 4686.

5. Hachmann, N.P., Miller, J., Collier, A.Y., Ventura, J.D., Yu, J., Rowe, M., Bondzie, E.A., Powers, O., Surve, N., Hall, K. and Barouch, D.H. (2022). Neutralization Escape by SARS-CoV- 2 Omicron Subvariants BA.2.12.1, BA.4, and BA.5. N Engl J Med 387, 86–88.

6. Al-Aly, Z., Bowe, B. and Xie, Y. (2022). Long COVID after breakthrough SARS-CoV-2 infection. Nat Med 28, 1461–1467.

7. Brannock, M.D., Chew, R.F., Preiss, A.J., Hadley, E.C., Redfield, S., McMurry, J.A., Leese, P.J., Girvin, A.T., Crosskey, M., Zhou, A.G., Moffitt, R.A., Funk, M.J., Pfaff, E.R., Haendel, M.A., Chute, C.G., N3C and Consortia, R. (2023). Long COVID risk and pre-COVID vaccination in an EHR-based cohort study from the RECOVER program. Nat Commun 14, 2914.

8. Kotfis, K., Williams Roberson, S., Wilson, J.E., Dabrowski, W., Pun, B.T. and Ely, E.W. (2020). COVID-19: ICU delirium management during SARS-CoV-2 pandemic. Crit Care 24, 176.

9. Mao, L., Jin, H., Wang, M., Hu, Y., Chen, S., He, Q., Chang, J., Hong, C., Zhou, Y., Wang, D., Miao, X., Li, Y. and Hu, B. (2020). Neurologic Manifestations of Hospitalized Patients With Coronavirus Disease 2019 in Wuhan, China. JAMA Neurol 77, 683–690.

10. DeKosky, S.T., Kochanek, P.M., Valadka, A.B., Clark, R.S.B., Chou, S.H., Au, A.K., Horvat, C., Jha, R.M., Mannix, R., Wisniewski, S.R., Wintermark, M., Rowell, S.E., Welch, R.D., Lewis, L., House, S., Tanzi, R.E., Smith, D.R., Vittor, A.Y., Denslow, N.D., Davis, M.D., Glushakova, O.Y. and Hayes, R.L. (2021). Blood Biomarkers for Detection of Brain Injury in COVID-19 Patients. J Neurotrauma 38, 1–43.

11. Lucchese, G., Vogelgesang, A., Boesl, F., Raafat, D., Holtfreter, S., Broker, B.M., Stufano, A., Fleischmann, R., Pruss, H., Franke, C. and Floel, A. (2022). Anti-neuronal antibodies against brainstem antigens are associated with COVID-19. EBioMedicine 83, 104211.

12. Heneka, M.T., Golenbock, D., Latz, E., Morgan, D. and Brown, R. (2020). Immediate and long-term consequences of COVID-19 infections for the development of neurological disease. Alzheimers Res Ther 12, 69.

13. Lee, M.H., Perl, D.P., Nair, G., Li, W., Maric, D., Murray, H., Dodd, S.J., Koretsky, A.P., Watts, J.A., Cheung, V., Masliah, E., Horkayne-Szakaly, I., Jones, R., Stram, M.N., Moncur, J., Hefti, M., Folkerth, R.D. and Nath, A. (2021). Microvascular Injury in the Brains of Patients with Covid-19. N Engl J Med 384, 481–483.

14. Stein, S.R., Ramelli, S.C., Grazioli, A., Chung, J.Y., Singh, M., Yinda, C.K., Winkler, C.W., Sun, J., Dickey, J.M., Ylaya, K., Ko, S.H., Platt, A.P., Burbelo, P.D., Quezado, M., Pittaluga, S., Purcell, M., Munster, V.J., Belinky, F., Ramos-Benitez, M.J., Boritz, E.A., Lach, I.A., Herr, D.L., Rabin, J., Saharia, K.K., Madathil, R.J., Tabatabai, A., Soherwardi, S., McCurdy, M.T., Consortium, N.C.-A., Peterson, K.E., Cohen, J.I., de Wit, E., Vannella, K.M., Hewitt, S.M., Kleiner, D.E. and Chertow, D.S. (2022). SARS-CoV-2 infection and persistence in the human body and brain at autopsy. Nature 612, 758–763.

15. Crunfli, F., Carregari, V.C., Veras, F.P., Silva, L.S., Nogueira, M.H., Antunes, A., Vendramini, P.H., Valenca, A.G.F., Brandao-Teles, C., Zuccoli, G.D.S., Reis-de-Oliveira, G., Silva-Costa, L.C., Saia-Cereda, V.M., Smith, B.J., Codo, A.C., de Souza, G.F., Muraro, S.P., Parise, P.L., Toledo-Teixeira, D.A., Santos de Castro, I.M., Melo, B.M., Almeida, G.M., Firmino, E.M.S., Paiva, I.M., Silva, B.M.S., Guimaraes, R.M., Mendes, N.D., Ludwig, R.L., Ruiz, G.P., Knittel, T.L., Davanzo, G.G., Gerhardt, J.A., Rodrigues, P.B., Forato, J., Amorim, M.R., Brunetti, N.S., Martini, M.C., Benatti, M.N., Batah, S.S., Siyuan, L., Joao, R.B., Aventurato, I.K., Rabelo de Brito, M., Mendes, M.J., da Costa, B.A., Alvim, M.K.M., da Silva Junior, J.R., Damiao, L.L., de Sousa, I.M.P., da Rocha, E.D., Goncalves, S.M., Lopes da Silva, L.H., Bettini, V., Campos, B.M., Ludwig, G., Tavares, L.A., Pontelli, M.C., Viana, R.M.M., Martins, R.B., Vieira, A.S., Alves-Filho, J.C., Arruda, E., Podolsky-Gondim, G.G., Santos, M.V., Neder, L., Damasio, A., Rehen, S., Vinolo, M.A.R., Munhoz, C.D., Louzada-Junior, P., Oliveira, R.D., Cunha, F.Q., Nakaya, H.I., Mauad, T., Duarte-Neto, A.N., Ferraz da Silva, L.F., Dolhnikoff, M., Saldiva, P.H.N., Farias, A.S., Cendes, F., Moraes-Vieira, P.M.M., Fabro, A.T., Sebollela, A., Proenca-Modena, J.L., Yasuda, C.L., Mori, M.A., Cunha, T.M. and Martins-de-Souza, D. (2022). Morphological, cellular, and molecular basis of brain infection in COVID-19 patients. Proc Natl Acad Sci U S A 119, e2200960119.

16. Huang, S. and Fishell, G. (2022). In SARS-CoV-2, astrocytes are in it for the long haul. Proc Natl Acad Sci U S A 119, e2209130119.

17. Serrano, G.E., Walker, J.E., Arce, R., Glass, M.J., Vargas, D., Sue, L.I., Intorcia, A.J., Nelson, C.M., Oliver, J., Papa, J., Russell, A., Suszczewicz, K.E., Borja, C.I., Belden, C., Goldfarb, D., Shprecher, D., Atri, A., Adler, C.H., Shill, H.A., Driver-Dunckley, E., Mehta, S.H., Readhead, B., Huentelman, M.J., Peters, J.L., Alevritis, E., Bimi, C., Mizgerd, J.P., Reiman, E.M., Montine, T.J., Desforges, M., Zehnder, J.L., Sahoo, M.K., Zhang, H., Solis, D., Pinsky, B.A., Deture, M., Dickson, D.W. and Beach, T.G. (2021). Mapping of SARS-CoV-2 Brain Invasion and Histopathology in COVID-19 Disease. medRxiv.

18. Taquet, M., Skorniewska, Z., Hampshire, A., Chalmers, J.D., Ho, L.P., Horsley, A., Marks, M., Poinasamy, K., Raman, B., Leavy, O.C., Richardson, M., Elneima, O., McAuley, H.J.C., Shikotra, A., Singapuri, A., Sereno, M., Saunders, R.M., Harris, V.C., Houchen-Wolloff, L., Greening, N.J., Mansoori, P., Harrison, E.M., Docherty, A.B., Lone, N.I., Quint, J., Sattar, N., Brightling, C.E., Wain, L.V., Evans, R.E., Geddes, J.R., Harrison, P.J. and Group, P.-C.S.C. (2023). Acute blood biomarker profiles predict cognitive deficits 6 and 12 months after COVID- 19 hospitalization. Nat Med 29, 2498–2508.

19. Poyiadji, N., Shahin, G., Noujaim, D., Stone, M., Patel, S. and Griffith, B. (2020). COVID- 19-associated Acute Hemorrhagic Necrotizing Encephalopathy: Imaging Features. Radiology 296, E119–E120.

20. Reichard, R.R., Kashani, K.B., Boire, N.A., Constantopoulos, E., Guo, Y. and Lucchinetti, C.F. (2020). Neuropathology of COVID-19: a spectrum of vascular and acute disseminated encephalomyelitis (ADEM)-like pathology. Acta Neuropathol 140, 1–6.

21. 21. Mavrikaki, M., Lee, J.D., Solomon, I.H. and Slack, F.J. (2021). Severe COVID-19 induces molecular signatures of aging in the human brain. medRxiv.

22. Hampshire, A., Chatfield, D.A., AM M.P., Jolly, A., Trender, W., Hellyer, P.J., Giovane, M.D., Newcombe, V.F.J., Outtrim, J.G., Warne, B., Bhatti, J., Pointon, L., Elmer, A., Sithole, N., Bradley, J., Kingston, N., Sawcer, S.J., Bullmore, E.T., Rowe, J.B., Menon, D.K., Cambridge NeuroCovid Group, t.N.C.-B. and Cambridge, N.C.R.F. (2022). Multivariate profile and acute-phase correlates of cognitive deficits in a COVID-19 hospitalised cohort. EClinicalMedicine 47, 101417.

23. Zarifkar, P., Peinkhofer, C., Benros, M.E. and Kondziella, D. (2022). Frequency of Neurological Diseases After COVID-19, Influenza A/B and Bacterial Pneumonia. Front Neurol 13, 904796.

24. Wang, L., Davis, P.B., Volkow, N.D., Berger, N.A., Kaelber, D.C. and Xu, R. (2022). Association of COVID-19 with New-Onset Alzheimer’s Disease. J Alzheimers Dis 89, 411–414.

25. Johansson, M.A., Quandelacy, T.M., Kada, S., Prasad, P.V., Steele, M., Brooks, J.T., Slayton, R.B., Biggerstaff, M. and Butler, J.C. (2021). SARS-CoV-2 Transmission From People Without COVID-19 Symptoms. JAMA Netw Open 4, e2035057.

26. Malkova, A., Kudryavtsev, I., Starshinova, A., Kudlay, D., Zinchenko, Y., Glushkova, A., Yablonskiy, P. and Shoenfeld, Y. (2021). Post COVID-19 Syndrome in Patients with Asymptomatic/Mild Form. Pathogens 10.

27. Ledford, H. (2022). How common is long COVID? Why studies give different answers. Nature 606, 852–853.

28. Zhang, H., Zang, C., Xu, Z., Zhang, Y., Xu, J., Bian, J., Morozyuk, D., Khullar, D., Zhang, Y., Nordvig, A.S., Schenck, E.J., Shenkman, E.A., Rothman, R.L., Block, J.P., Lyman, K., Weiner, M.G., Carton, T.W., Wang, F. and Kaushal, R. (2023). Data-driven identification of post-acute SARS-CoV-2 infection subphenotypes. Nat Med 29, 226–235.

29. 29. Ballering, A.V., van Zon, S.K.R., Olde Hartman, T.C., Rosmalen, J.G.M. and Lifelines Corona Research, I. (2022). Persistence of somatic symptoms after COVID-19 in the Netherlands: an observational cohort study. Lancet 400, 452–461.

30. Hastie, C.E., Lowe, D.J., McAuley, A., Winter, A.J., Mills, N.L., Black, C., Scott, J.T., O’Donnell, C.A., Blane, D.N., Browne, S., Ibbotson, T.R. and Pell, J.P. (2022). Outcomes among confirmed cases and a matched comparison group in the Long-COVID in Scotland study. Nat Commun 13, 5663.

31. Qasmieh, S.A., Robertson, M.M., Teasdale, C.A., Kulkarni, S.G., Jones, H.E., McNairy, M., Borrell, L.N. and Nash, D. (2023). The prevalence of SARS-CoV-2 infection and long COVID in U.S. adults during the BA.4/BA.5 surge, June-July 2022. Prev Med 169, 107461.

32. Xu, E., Xie, Y. and Al-Aly, Z. (2022). Long-term neurologic outcomes of COVID-19. Nat Med 28, 2406–2415.

33. Statistics, C.N.C.f.H. (2022). Nearly One in Five American Adults Who Have Had COVID-19 Still Have "Long COVID": Online: https://www.cdc.gov/nchs/pressroom/nchs_press_releases/2022/20220622.htm.

34. Antonelli, M., Pujol, J.C., Spector, T.D., Ourselin, S. and Steves, C.J. (2022). Risk of long COVID associated with delta versus omicron variants of SARS-CoV-2. Lancet 399, 2263–2264.

35. Kahlert, C.R., Strahm, C., Güsewell, S., Cusini, A., Brucher, A., Goppel, S., Möller, J.C., Ortner, M., Ruetti, M., Stocker, R., Vuichard-Gysin, D., Besold, U., McGeer, A., Risch, L., Friedl, A., Schlegel, M., Vernazza, P., Kuster, S.P. and Kohler, P. (2022). Association of viral variant and vaccination status with the occurrence of symptoms compatible with post-acute sequelae after primary SARS-CoV-2 infection. medRxiv, 2022.2010.2021.22281349.

36. Choutka, J., Jansari, V., Hornig, M. and Iwasaki, A. (2022). Unexplained post-acute infection syndromes. Nat Med 28, 911–923.

37. Damiano, R.F., Guedes, B.F., de Rocca, C.C., de Padua Serafim, A., Castro, L.H.M., Munhoz, C.D., Nitrini, R., Filho, G.B., Miguel, E.C., Lucchetti, G. and Forlenza, O. (2022). Cognitive decline following acute viral infections: literature review and projections for post-COVID-19. European archives of psychiatry and clinical neuroscience 272, 139–154.

38. Abraham, G.R., Kuc, R.E., Althage, M., Greasley, P.J., Ambery, P., Maguire, J.J., Wilkinson, I.B., Hoole, S.P., Cheriyan, J. and Davenport, A.P. (2022). Endothelin-1 is increased in the plasma of patients hospitalised with Covid-19. J Mol Cell Cardiol 167, 92–96.

39. Fernandez-Castaneda, A., Lu, P., Geraghty, A.C., Song, E., Lee, M.H., Wood, J., O’Dea, M.R., Dutton, S., Shamardani, K., Nwangwu, K., Mancusi, R., Yalcin, B., Taylor, K.R., Acosta-Alvarez, L., Malacon, K., Keough, M.B., Ni, L., Woo, P.J., Contreras-Esquivel, D., Toland, A.M.S., Gehlhausen, J.R., Klein, J., Takahashi, T., Silva, J., Israelow, B., Lucas, C., Mao, T., Pena-Hernandez, M.A., Tabachnikova, A., Homer, R.J., Tabacof, L., Tosto-Mancuso, J., Breyman, E., Kontorovich, A., McCarthy, D., Quezado, M., Vogel, H., Hefti, M.M., Perl, D.P., Liddelow, S., Folkerth, R., Putrino, D., Nath, A., Iwasaki, A. and Monje, M. (2022). Mild respiratory COVID can cause multi-lineage neural cell and myelin dysregulation. Cell 185, 2452–2468 e2416.

40. Frontera, J.A., Yang, D., Lewis, A., Patel, P., Medicherla, C., Arena, V., Fang, T., Andino, A., Snyder, T., Madhavan, M., Gratch, D., Fuchs, B., Dessy, A., Canizares, M., Jauregui, R., Thomas, B., Bauman, K., Olivera, A., Bhagat, D., Sonson, M., Park, G., Stainman, R., Sunwoo, B., Talmasov, D., Tamimi, M., Zhu, Y., Rosenthal, J., Dygert, L., Ristic, M., Ishii, H., Valdes, E., Omari, M., Gurin, L., Huang, J., Czeisler, B.M., Kahn, D.E., Zhou, T., Lin, J., Lord, A.S., Melmed, K., Meropol, S., Troxel, A.B., Petkova, E., Wisniewski, T., Balcer, L., Morrison, C., Yaghi, S. and Galetta, S. (2021). A prospective study of long-term outcomes among hospitalized COVID-19 patients with and without neurological complications. J Neurol Sci 426, 117486.

41. Klein, J., Wood, J., Jaycox, J., Lu, P., Dhodapkar, R.M., Gehlhausen, J.R., Tabachnikova, A., Tabacof, L., Malik, A.A., Kamath, K., Greene, K., Monteiro, V.S., Pena-Hernandez, M., Mao, T., Bhattacharjee, B., Takahashi, T., Lucas, C., Silva, J., McCarthy, D., Breyman, E., Tosto-Mancuso, J., Dai, Y., Perotti, E., Akduman, K., Tzeng, T.J., Xu, L., Yildirim, I., Krumholz, H.M., Shon, J., Medzhitov, R., Omer, S.B., van Dijk, D., Ring, A.M., Putrino, D. and Iwasaki, A. (2022). Distinguishing features of Long COVID identified through immune profiling. medRxiv.

42. Masvekar, R.R., Kosa, P., Jin, K., Dobbs, K., Stack, M.A., Castagnoli, R., Quaresima, V., Su, H.C., Imberti, L., Notarangelo, L.D. and Bielekova, B. (2022). Prognostic value of serum/plasma neurofilament light chain for COVID-19-associated mortality. Ann Clin Transl Neurol 9, 622–632.

43. Needham, E.J., Ren, A.L., Digby, R.J., Norton, E.J., Ebrahimi, S., Outtrim, J.G., Chatfield, D.A., Manktelow, A.E., Leibowitz, M.M., Newcombe, V.F.J., Doffinger, R., Barcenas-Morales, G., Fonseca, C., Taussig, M.J., Burnstein, R.M., Samanta, R.J., Dunai, C., Sithole, N., Ashton, N.J., Zetterberg, H., Gisslen, M., Eden, A., Marklund, E., Openshaw, P.J.M., Dunning, J., Griffiths, M.J., Cavanagh, J., Breen, G., Irani, S.R., Elmer, A., Kingston, N., Summers, C., Bradley, J.R., Taams, L.S., Michael, B.D., Bullmore, E.T., Smith, K.G.C., Lyons, P.A., Coles, A.J., Menon, D.K., Cambridge Neuro, C.G., Collaboration, C.-N.C.-B. and Cambridge, N.C.R.F. (2022). Brain injury in COVID-19 is associated with dysregulated innate and adaptive immune responses. Brain 145, 4097–4107.

44. Peluso, M.J., Deeks, S.G., Mustapic, M., Kapogiannis, D., Henrich, T.J., Lu, S., Goldberg, S.A., Hoh, R., Chen, J.Y., Martinez, E.O., Kelly, J.D., Martin, J.N. and Goetzl, E.J. (2022). SARS-CoV-2 and Mitochondrial Proteins in Neural-Derived Exosomes of COVID-19. Ann Neurol 91, 772–781.

45. Swank, Z., Senussi, Y., Manickas-Hill, Z., Yu, X.G., Li, J.Z., Alter, G. and Walt, D.R. (2023). Persistent Circulating Severe Acute Respiratory Syndrome Coronavirus 2 Spike Is Associated With Post-acute Coronavirus Disease 2019 Sequelae. Clin Infect Dis 76, e487–e490.

46. Frontera, J.A., Boutajangout, A., Masurkar, A.V., Betensky, R.A., Ge, Y., Vedvyas, A., Debure, L., Moreira, A., Lewis, A., Huang, J., Thawani, S., Balcer, L., Galetta, S. and Wisniewski, T. (2022). Comparison of serum neurodegenerative biomarkers among hospitalized COVID-19 patients versus non-COVID subjects with normal cognition, mild cognitive impairment, or Alzheimer’s dementia. Alzheimers Dement 18, 899–910.

47. Peluso, M.J., Sans, H.M., Forman, C.A., Nylander, A.N., Ho, H.E., Lu, S., Goldberg, S.A., Hoh, R., Tai, V., Munter, S.E., Chenna, A., Yee, B.C., Winslow, J.W., Petropoulos, C.J., Martin, J.N., Kelly, J.D., Durstenfeld, M.S., Hsue, P.Y., Hunt, P.W., Greene, M., Chow, F.C., Hellmuth, J., Henrich, T.J., Glidden, D.V. and Deeks, S.G. (2022). Plasma Markers of Neurologic Injury and Inflammation in People With Self-Reported Neurologic Postacute Sequelae of SARS-CoV-2 Infection. Neurol Neuroimmunol Neuroinflamm 9.

48. Bazarian, J.J., Biberthaler, P., Welch, R.D., Lewis, L.M., Barzo, P., Bogner-Flatz, V., Gunnar Brolinson, P., Buki, A., Chen, J.Y., Christenson, R.H., Hack, D., Huff, J.S., Johar, S., Jordan, J.D., Leidel, B.A., Lindner, T., Ludington, E., Okonkwo, D.O., Ornato, J., Peacock, W.F., Schmidt, K., Tyndall, J.A., Vossough, A. and Jagoda, A.S. (2018). Serum GFAP and UCH-L1 for prediction of absence of intracranial injuries on head CT (ALERT-TBI): a multicentre observational study. Lancet Neurol 17, 782–789.

49. Korley, F.K., Jain, S., Sun, X., Puccio, A.M., Yue, J.K., Gardner, R.C., Wang, K.K.W., Okonkwo, D.O., Yuh, E.L., Mukherjee, P., Nelson, L.D., Taylor, S.R., Markowitz, A.J., Diaz-Arrastia, R., Manley, G.T. and Investigators, T.-T.S. (2022). Prognostic value of day-of-injury plasma GFAP and UCH-L1 concentrations for predicting functional recovery after traumatic brain injury in patients from the US TRACK-TBI cohort: an observational cohort study. Lancet Neurol 21, 803–813.

50. Lewis, L.M., Papa, L., Bazarian, J.J., Weber, A., Howard, R. and Welch, R.D. (2020). Biomarkers May Predict Unfavorable Neurological Outcome after Mild Traumatic Brain Injury. J Neurotrauma 37, 2624–2631.

51. 51. GraphPad Software, L.L.C. (2023). GraphPad Prism Statistics Guide - Principles of Statistics: https://www.graphpad.com/guides/prism/latest/statistics/stat_---_principles_of_statistics_-.htm

52. NIST/SEMATECH e-Handbook of Statistical Method: https://www.itl.nist.gov/div898/handbook/

53. Luger, S., Jaeger, H.S., Dixon, J., Bohmann, F.O., Schaefer, J., Richieri, S.P., Larsen, K., Hov, M.R., Bache, K.G., Foerch, C. and Group, B.F.I.S. (2020). Diagnostic Accuracy of Glial Fibrillary Acidic Protein and Ubiquitin Carboxy-Terminal Hydrolase-L1 Serum Concentrations for Differentiating Acute Intracerebral Hemorrhage from Ischemic Stroke. Neurocrit Care 33, 39–48.

54. Papa, L. and Wang, K.K.W. (2017). Raising the Bar for Traumatic Brain Injury Biomarker Research: Methods Make a Difference. J Neurotrauma 34, 2187–2189.

55. 55. Andrews, M.G., Mukhtar, T., Eze, U.C., Simoneau, C.R., Perez, Y., Mostajo-Radji, M.A., Wang, S., Velmeshev, D., Salma, J., Kumar, G.R., Pollen, A.A., Crouch, E.E., Ott, M. and Kriegstein, A.R. (2021). Tropism of SARS-CoV-2 for Developing Human Cortical Astrocytes. bioRxiv.

56. Levine, K.S., Leonard, H.L., Blauwendraat, C., Iwaki, H., Johnson, N., Bandres-Ciga, S., Ferrucci, L., Faghri, F., Singleton, A.B. and Nalls, M.A. (2023). Virus exposure and neurodegenerative disease risk across national biobanks. Neuron 111, 1086–1093 e1082.

57. 57. Carneiro, V.C.S., Alves-Leon, S.V., Sarmento, D.J.S., Coelho, W., Moreira, O.D.C., Salvio, A.L., Ramos, C.H.F., Ramos Filho, C.H.F., Marques, C.A.B., da Costa Goncalves, J.P., Leon, L.A.A. and de Paula, V.S. (2022). Herpesvirus and neurological manifestations in patients with severe coronavirus disease. Virology journal 19, 101.

