## Supplemental Table 1 for "An FDA-approved assay platform can detect biomarkers of neuronal and glial injury in the blood of COVID-19 patients"

**<u>Supplemental Table 1</u>: Demographics, Clinical Characteristics and Biomarker Values in the COVID-19-positive Patients Enrolled in the Study between March, 27 2020 and April, 27 2020.**

| Subject ID | Age range (years) | Sex | Neurologic injury/changes | Supplemental Oxygen Requirement | Neuroimaging Chart Review (Feb 2023) | Other information | Day of 1st sample collection | Peak UCH-L1 | Peak GFAP | Initial UCH-L1 | Initial GFAP |
|---|---|---|---|---|---|---|---|---|---|---|---|
| 005 | 61-65 | M | Encephalopathy after extubation, but no imaging | Intubated | No brain imaging | Died in hospital | 7 | 271.6 (D7)* | 57.1 (D7)* | 271.6 (D7)* | 57.1 (D7)* |
| 007 | 51-55 | M | Had multiple episode of unilateral weakness while in hospital which resolved, had Brain MRI after discharge showing diffuse microhemorrhages | Intubated, ECMO | Negative Head CT 18 days after enrollment, outpatient Brain MRI was 2.5 months after enrollment | | 3 | 825.6 (D14) | 18.1 (D14) | 687.3 (D3) | 7.4 (D3) |
| 011 | 66-69 | M | None | Intubated | No brain imaging | | 3 | 355.5 (D7) | 31.4 (D3) | 262.3 (D3) | 31.4 (D3) |
| 012 | 61-65 | M | None | Required 2L NC | No brain imaging | | 28 | 409.6 (D28)* | 15.9 (D28)* | 409.6 (D28)* | 15.9 (D28)* |
| 013 | 76-80 | M | Had extremity weakness in hospital, CT with acute/subacute hypoattenuation | Intubated | Head CT was 18 days after enrollment | | 3 | 619.5 (D14) | 53.9 (D0) | 332.6 (D0) | 53.9 (D0) |
| 014 | 66-70 | M | None | Intubated, ARDS, proned | No brain imaging | | 0 | 540.3 (D7) | 40.2 (D0) | **243.3 (D0)** | **40.2 (D0)** |
| 021 | 66-70 | F | Encephalopathy after extubation, persistent cognitive deficits, CT with microvascular ischemic disease and intracranial atherosclerosis | Intubated | Head CT was 28 days after enrollment | | 0 | 813 (D0) | 212.7 (D3) | 813 (D0) | 174.8 (D0) |
| 025 | 61-65 | M | None | Required 3L NC | No brain imaging | | 0 | 158.6 (D0)* | 62.3 (D0)* | 158.6 (D0)* | 62.3 (D0)* |
| 027 | 56-60 | M | Seizure and then coded in hospital, no imaging | Required 3L NC, then 6L NC | No brain imaging | Died in hospital | 0 | 352.8 (D0) | 9.2 (D0) | 352.8 (D0) | 9.2 (D0) |

| | | | | | | | | | | | |
|---|---|---|---|---|---|---|---|---|---|---|---|
| 031 | 86-90 | F | None | Required initially 2L NC, max 10L NC | Head CT 3 months after enrollment with age-indeterminant locunar infarct | | 0 | 509.8 (D7) | 112.7 (D14) | 269.7 (D0) | 35.2 (D0) |
| 038 | 61-65 | F | None | Required 4L NC | Negative head CT after a fall 14 days after enrollment | | 3 | 740.5 (D3)* | 133.1 (D3)* | 740.5 (D3)* | 133.1 (D3)* |
| 039 | 71-75 | F | AMS, neg head CT, neg EEG | Required 3L NC | Had negative head CTs 3 days, 5 days, and 15 days after enrollment | | 7 | 253.1 (D7)* | 60.3 (D7)* | 253.1 (D7)* | 60.3 (D7)* |
| 041 | 71-75 | M | AMS, neg head CT | Required 6L NC | Head CT was on day of enrollment | | 0 | 462.6 (D7) | 49.4 (D7) | 186.2 (D0) | 30.1 (D0) |
| 042 | 61-65 | M | AMS, neg head CT | None | Head CT was the day before enrollment | | 0 | 1010.7 (D7) | 57.4 (D3) | 227.1 (D0) | 41.5 (D0) |
| 047 | 66-70 | M | AMS, no imaging | Required high flow NC, did not intubate as DNI | No brain imaging | | 0 | 756.6 (D3) | 54.3 (D0) | 554.8 (D0) | 54.3 (D0) |
| 051 | 66-70 | F | None | Required 4L NC | No brain imaging | | 0 | 178.4 (D0)* | 9.2 (D0)* | 178.4 (D0)* | 9.2 (D0)* |
| 052 | 51-55 | M | None | Required 3L NC | No brain imaging | | 0 | 202.3 (D0)* | 121.5 (D0)* | 202.3 (D0)* | 121.5 (D0)* |
| 055 | 76-80 | M | AMS, neg head CT | None | Had negative head CTs 2 days and 18 days after enrollment | | 0 | 615 (D3) | 102.1 (D3) | 475.3 (D0) | 89.6 (D0) |
| 059 | 51-55 | M | None | Required 2L NC | No brain imaging | | 0 | 248.3 (D0)* | 13.8 (D0)* | 248.3 (D0)* | 13.8 (D0)* |
| 060 | 51-55 | F | None | Required 2L NC | No brain imaging | | 3 | 180.2 (D3)* | 46.8 (D3)* | 180.2 (D3)* | 46.8 (D3)* |
| 061 | 56-60 | M | None | Required 4L NC | No brain imaging | | 0 | 253.3 (D0) | 9.8 (D3) | 253.3 (D0) | 6.6 (D0) |

| | | | | | | | | | | | |
|---|---|---|---|---|---|---|---|---|---|---|---|
| 064 | 61-65 | M | Presented with acute right subcortical ischemic stroke, confirmed on brain MRI, also found to have small L SDH on this MRI, encephalopathy developed while inpatient, neg LP | None | Had negative head CT on the day of enrollment and the day after enrollment. Brain MRI which showed acute CVA and small SDH was on day after enrollment. Additional head CT 3 days after enrollment showed SDH. Additional head CTs 6 days and 21 days after enrollment showed the evolving CVA and SDH. | | 7 | 417.1 (D7)* | 35.1 (D7)* | 417.1 (D7)* | 35.1 (D7)* |
| 080 | 86-90 | M | Confusion | Intubated | Negative head CT was on day of enrollment. Head CT 21 days after enrollment showed a acute vs. subacute right occipital lob infarct. | | 0 | 8106 (D0) | 382.4 (D0) | 8106 (D0) | 382.4 (D0) |
| 085 | 76-80 | M | Confusion | Yes | Negative head CT was on day before enrollment. | | 3 | 712.5 (D3) | 69.6 (D3) | 712.5 (D3) | 69.6 (D3) |
| 090 | 61-65 | M | Loss of smell | No | No brain imaging | | 0 | 311.1 (D0)* | 92.6 (D0)* | 311.1 (D0)* | 92.6 (D0)* |
| 092 | 71-75 | M | ICH | No | Head CT day before enrollment showed small ICH in right corona radiata and old right MCA and left vertebral artery occlusion. On day before enrollment, subject presented after being found down with altered mental status and new left hemiparesis and neglect. | Old R MCA CVA | 0 | 456.5 (D14)* | 55.4 (D14)* | 456.5 (D14)* | 55.4 (D14)* |
| 101 | 36-40 | F | Loss of smell | Mild hypoxemia | No brain imaging | Admitted 9 days after the first positive test. | 0 | 159.7 (D0)* | <10 (D0)* | 159.7 (D0)* | <10 (D0)* |

| | | | | | | | | | | | |
|---|---|---|---|---|---|---|---|---|---|---|---|
| 107 | 61-65 | M | Headache | Moderate | Had negative head CT on day of enrollment Acute internal capsule CVA was seen on head CT 2 months after enrollment. | Elevated d-dimer. | 0 | 445.6 (D3) | 19.1 (D0) | 400.1 (D0) | 19.1 (D0) |
| 116 | 81-85 | M | Aphasia, weakness | Severe hypoxemia | Head CT the day before enrollment showed old infarcts. | Old CVAs | 0 | 3696.6 (D3) | 161.9 (D3) | 863.4 (D0) | 61.6 (D0) |
| 119 | 36-40 | F | Unresponsive | Severe hypoxemia | Head CT the day before enrollment showed diffuse cerebral edema consistent with hypoxic ischemic injury. | Respiratory/ cardiac arrest. CT diffuse injury | 0 | 1635.5 (D0)* | 121 (D0)* | 1635.5 (D0)* | 121 (D0)* |
| 120 | 56-60 | F | Unresponsive | Severe hypoxemia | MRI with cerebellar punctate microhemorrhages was 1.5 months after enrollment. | | 0 | 489.7 (D0) | 99.8 (D3) | 489.7 (D0) | 78.9 (D0) |
| 128 | 26-30 | M | AMS (delayed) | | Had negative head CT 3 days after enrollment | Admitted and discharged on the same day and readmitted on the next day | 0 | 348 (D0)* | 8.5 (D0)* | 348 (D0)* | 8.5 (D0)* |
| 131 | 51-55 | M | Decreased taste | Moderate hypoxemia | No brain imaging | Admitted 9 days after the first positive test. | 3 | 256.3 (D3) | 25.1 (D7) | 256.3 (D3) | 21.6 (D3) |

*Designates it is the only sample obtained; in columns representing initial and peak biomarker concentrations, the values are in pg/ml and (D) signifies the day of sample collection.

Abbreviations: AMS- altered mental status, ARDS- acute respiratory distress syndrome, CT- computed tomography, CVA- cerebral vascular accident, DNI- do not intubate, ECMO- extracorporeal membrane oxygenation, EEG- electroencephalogram, GFAP- glial fibrillary acidic protein, ICH- intracranial hemorrhage, MCA- middle cerebral artery, MRI- magnetic resonance imaging, NC- nasal cannula, SDH- subdural hematoma, UCH-L1- Ubiquitin C-terminal hydrolase-L1.
